# Meteorological factors and domestic new cases of coronavirus disease (COVID-19) in nine Asian cities: A time-series analysis

**DOI:** 10.1101/2020.04.15.20066613

**Authors:** Zonglin He, Yiqiao Chin, Jian Huang, Yi He, Babatunde O. Akinwunmi, Shinning Yu, Casper JP Zhang, Wai-kit Ming

## Abstract

**AIM:** To investigate the associations of meteorological factors and the daily new cases of coronavirus disease (COVID-19) in nine Asian cities.

**METHOD:** Pearson’s correlation and generalized additive modeling were performed to assess the relationships between daily new COVID-19 cases and meteorological factors (daily average temperature and relative humidity) with the most updated data currently available.

**RESULTS:** The Pearson correlation showed that daily new confirmed cases of COVID-19 were more correlated with the average temperature than with relative humidity. Daily new confirmed cases were negatively correlated with the average temperature in Beijing (r=-0.565, P<0.01), Shanghai (r=-0.471, P<0.01), and Guangzhou (r=-0.530, P<0.01), yet in contrast, positively correlated with that in Japan (r=0.441, P<0.01). In most of the cities (Shanghai, Guangzhou, Hong Kong, Seoul, Tokyo, and Kuala Lumpur), generalized additive modeling analysis showed the number of daily new confirmed cases was positively associated with both average temperature and relative humidity, especially in lagged 3d model, where a positive influence of temperature on the daily new confirmed cases was discerned in 5 cities except in Beijing, Wuhan, Korea, and Malaysia. Nevertheless, the results were inconsistent across cities and lagged time, suggesting meteorological factors were unlikely to greatly influence the COVID-19 epidemic.

**CONCLUSION:** The associations between meteorological factors and the number of COVID-19 daily cases are inconsistent across cities and lagged time. Large-scale public health measures and expanded regional research are still required until a vaccine becomes available and herd immunity is established.

**Significance statement:** With increasing COVID-19 cases across China and the world, and previous studies showing that meteorological factors may be associated with infectious disease transmission, the saying has it that when summer comes, the epidemic of COVID-19 may simultaneously fade away. We demonstrated the influence of meteorological factors on the daily domestic new cases of coronavirus disease (COVID-19) in nine Asian cities. And we found that the associations between meteorological factors and the number of COVID-19 daily cases are inconsistent across cities and time. We think this important topic may give better clues on prevention, management, and preparation for new events or new changes that could happen in the COVID-19 epidemiology in various geographical regions and as we move towards Summer.

## Introduction

In December 2019, several cases of pneumonia with an unknown etiology were reported in Wuhan, Hubei province, China (1). A novel strain of coronavirus was identified from the nasopharyngeal swab specimens of the infected, which was later designated as severe acute respiratory syndrome coronavirus 2 (SARS-CoV-2), and the resulting pneumonia as coronavirus disease (COVID-19). With the number of the infected soaring up, on 31^st^ January, 2020, the World Health Organization (WHO) has declared the outbreak of COVID-19 as a public health emergency of international concern (PHEIC) (2).

SARS-CoV-2, pertaining to the *Coronaviridae*, is an enveloped, single-stranded, positive-sense RNA virus, which is closely related to SARS-like coronaviruses, and based on the phylogenetic analysis, these coronaviruses have a common ancestor that resembles the bat coronavirus HKU9-1 (3, 4). Evidence has shown that the SARS-CoV-2 can transmit between human through respiratory droplets, fecal-oral route, direct contacts, and aerosols(5-7). Moreover, the long incubation period (1 to 14 days) increases the difficulty of controlling the COVID-19 outbreak. Studies have shown the mean incubation period was 5.1 days (ranged from 2.2 to 11.5 days, 95% confidence interval (CI): 4.5–5.8) (8), and another study estimated the mean incubation period to be 6.4 days (ranged from 2 to 14 days)(9). By March 28, 2020, more than 199 countries had confirmed cases, with more than 571,678 confirmed cases and 26,494 mortalities globally, among which 489,337 (85.6%) confirmed cases and 23,188 (87.5%) mortalities were reported outside of China (10).

Human respiratory pathogens (bacterial pathogens like pneumococcus and viruses like rubella and influenza) usually exhibit an annual pattern showing an increase in incidence during winter, and decrease during the summer (seasonality). Despite much data on influenza, respiratory syncytial virus, and the SARS 2003 outbreak following the pattern, it is difficult to predict whether COVID-19 would follow the trend and will be eliminated in the coming summer, as our understanding of the forces driving the seasonality of infectious diseases remains limited. Since influenza is a common viral disease, a proportion of the population already possesses some levels of immunity and when more patients recover, herd immunity constrain the transmission of the virus. The low or absent prevalence trend of SARS or MERS-CoV in the summer was also mentioned to be strongly relied on the use of effective therapeutic treatment and strict public health measures(11, 12). However, SARS-CoV-2 is a novel virus to humans, immunity to this ongoing virus pandemic is limited and an effective pharmaceutical therapy or vaccine prevention is yet to be found.

The seasonality in the outbreak of respiratory infectious diseases might be due to the seasonal variation of host physiology (susceptibility, individual immunity and herd immunity)(13), genetics(14), viral stability(15, 16) and infectivity(17-20), latent infectors constantly shedding viruses(21, 22), and the atmospheric dispersion and trans-ocean inter-continental migration (23-26), which are mainly driven by the meteorological factors, including the temperature and humidity (27). Geological features and latitude play a major role in forming a meteorological pattern. Lowen et al. assumed the seasonality of influenza and Respiratory Syncytial Virus (RSV) epidemics in temperate climates mainly attributable to the low absolute humidity (19), and the following factors associated with it, namely the low temperature, increased population, and low micronutrient levels (such as low vitamin D levels) (28, 29). During the winter in temperate zones, the temperature and humidity are low, characterized by dryness and coldness, and viruses are more easily to be transmitted via aerosols than direct or indirect contact, where the low temperature renders the virus viable and stable in aerosols, and on the surface. However, in the rainy seasons of tropical regions, where the weather is hot and wet, the aerosol transmission decreases but the transmission by direct contact increases(30); although the high temperature can decrease the stability of the virus and reduce the level of aerosolization of the viral droplet, with the increased temperature, the amount of virus deposited on surfaces increases(31).

Because of the relatively stable structure of the coronaviruses, the infectivity of the coronavirus would not be affected upon circumstances of a relative humidity >95% and a temperature of 28 - 33°C (31). For example, the transmission of SARS is more efficient in the temperature between 16 °C and 28°C, the widely use of air-conditioning also provides a shelter for the breeding and transmitting of SARS, where the virus is stable for 3 weeks at room temperature (31). SARS-CoV-2 has many similarities with the SARS-CoV-1, but whether meteorological factors influenced the viral transmission have not been established. Therefore, in the present study, we aimed to examine the relationship between meteorological factors and the number of COVID-19 daily cases in Asian cities at different latitudes.

## Method

### Meteorological data

We obtained daily meteorological data from the National Meteorological Information Center (http://cdc.cma.gov.cn/). The data were collected from meteorological stations in Beijing, Wuhan, Shanghai, Guangzhou, Hong Kong, Seoul, Tokyo, Kuala Lumpur and Singapore. Where data from the national meteorological stations were not available, we obtained data from the Timeanddate (https://www.timeanddate.com/). We retrieved the highest and lowest temperatures and humidity in four different quarters of the day and computed the average values of temperature and humidity for that day.

### Surveillance data of COVID-19

The daily number of domestic COVID-19 cases in five cities in China (namely Wuhan, Beijing, Shanghai, Guangzhou, and Hong Kong) and Singapore were obtained from the Ministry of Health in China and Singapore, respectively. Given that the daily number of domestic cases at the city level in Japan, Korea and Malaysia were not available from the corresponding Ministry of Health, as an alternative, we used the number of domestic cases at the national level in these three countries for analysis. The clinical criterion for the diagnosis of COVID-19 was based on the high-throughput sequencing or RT-PCR assay by nasopharyngeal swab.

### Statistical analysis

We evaluated the normality of the daily new cases and meteorological data by examining their skewness and kurtosis. We also estimated the Pearson correlation and covariances between the daily COVID-19 new cases and daily meteorological factors using STATA 14.0.

Generalized additive models (GAMs) with a Poisson family and logarithm link function were used to estimate the associations of daily COVID-19 new cases with average temperature and relative humidity. GAMs are useful for identifying exposure-response relationships from various types of data, particularly in exploring nonparametric relationships (32). The GAM analysis was performed in R software (version 3.6.0) using the package “mgcv”. We first established a basic temporal model for COVID-19 cases without including meteorological variables. To adjust for long-term trends and seasonality, we included penalized spline functions of time in the model. The degree of freedom (*df*) for time was optimized by minimizing of the absolute values of the partial autocorrelation function (PACF) of residuals for lags up to 30 days (33-36). Additionally, the selection of an optimal model was based on the lowest Akaike’s Information Criterion (AIC). Secondly, we built meteorological models based on the temporal models to account for the lagged effect of meteorological variables and the incubation period of COVID-19 (7, 37). Specifically, we examined the effect of meteorological variables with different time lags including one-day lag (Lag 1d), three-day lag (Lag 3d), five-day lag (Lag 5d), single-week lag (Lag 7d), and two-week lag (Lag 14) to capture immediate effects and lagged effects, respectively. Automated penalized splines were used to fit the association between the daily new cases and each of the meteorological variables. The date when the accumulated cases exceeded 30 in each city or country was selected as the inception of the date incorporated in the model to equalize the starting speed of outbreak and thus avoid miss-interpretation and overfitting.

To control for the autocorrelation, the model’s residuals were examined for serial correlation using PACF. Default plots showing the smooth components of a fitted GAM were produced. The percentage of deviance explained by each variable was calculated, which represents the scale of a linear predictor that contributes to the component smooth functions. At last, the graph of smooth components were grouped based on similar latitudes to mitigate the influence by latitude.

## Results

### Descriptive statistical results

The daily new cases of COVID-19 and meteorological data from January 20 to March 18, 2020 (61 days) in nine different cities are shown in Table 1. During the study period, the temperature in the five cities in China ranged from −9 to 26.25°C, with that ranging from −9 to 26°C in Beijing, from 0 to 22°C in Shanghai, from 8.75 to 26.25 °C in Guangzhou, from 1 to 21.75°C in Wuhan, and from 11.25 to 26.25°C in Hong Kong. The temperature in Singapore ranged from 25.5 to 32.75°C, in Seoul from −9.5 to 12.25°C, in Tokyo from 0.75 to 18.5°C, and in Kuala Lumpur from 24.5 to 33.5°C. Figures 1 and 2 show the distribution of daily COVID-19 new cases associated with average temperature and average relative humidity over time, respectively.

**Table 1.**
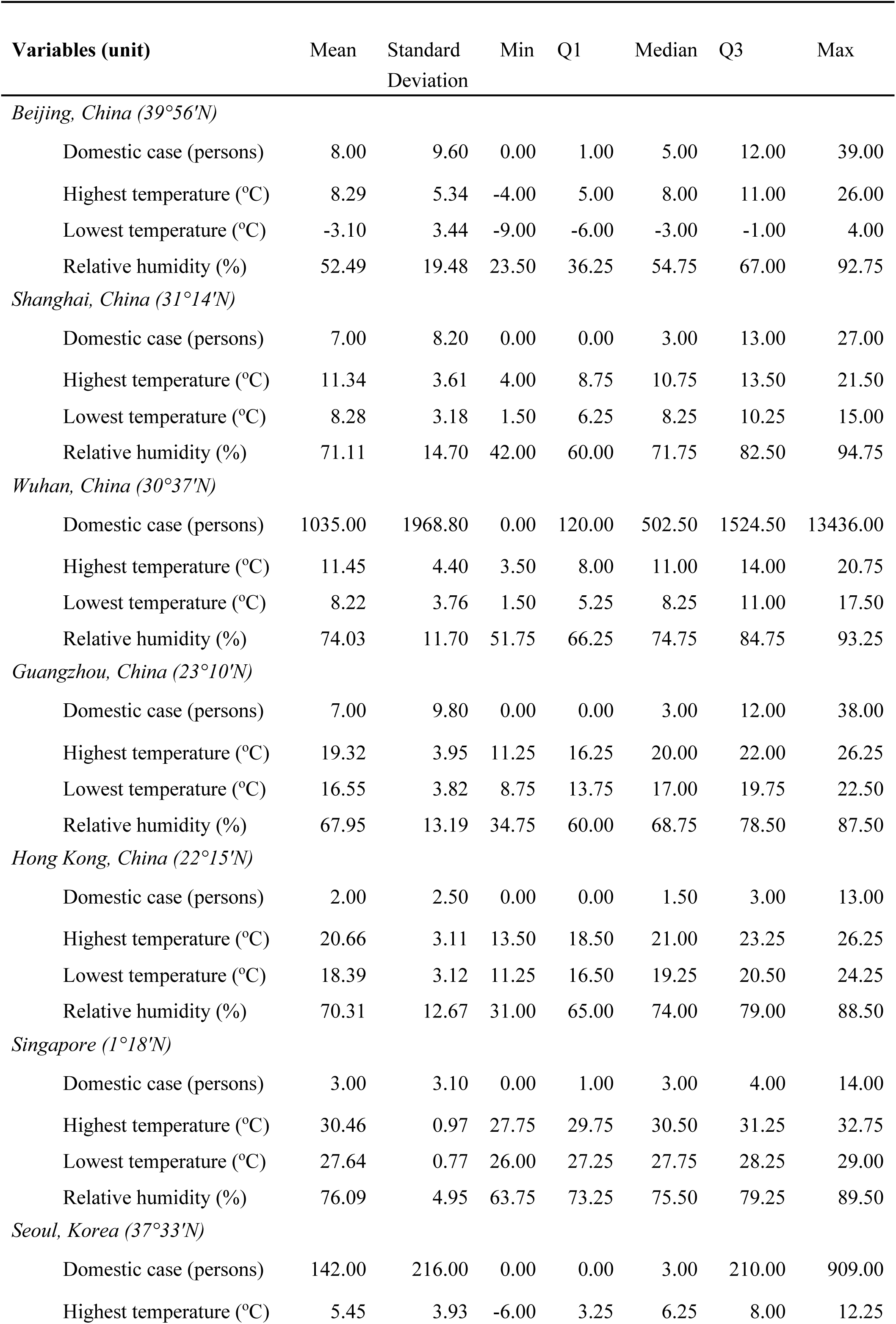
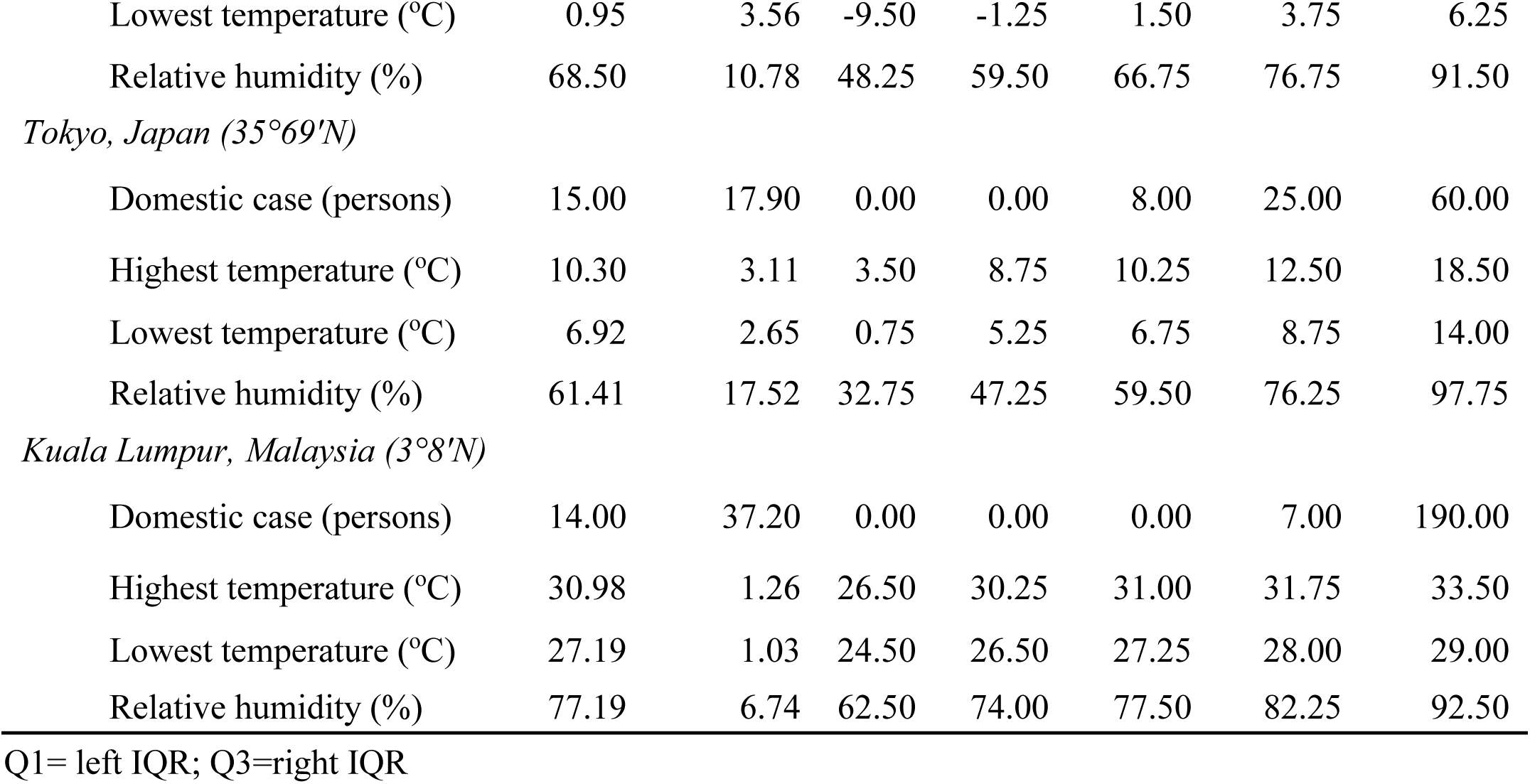
Descriptive Statistics for different Asian cities (with latitude)

**Figure 1.**
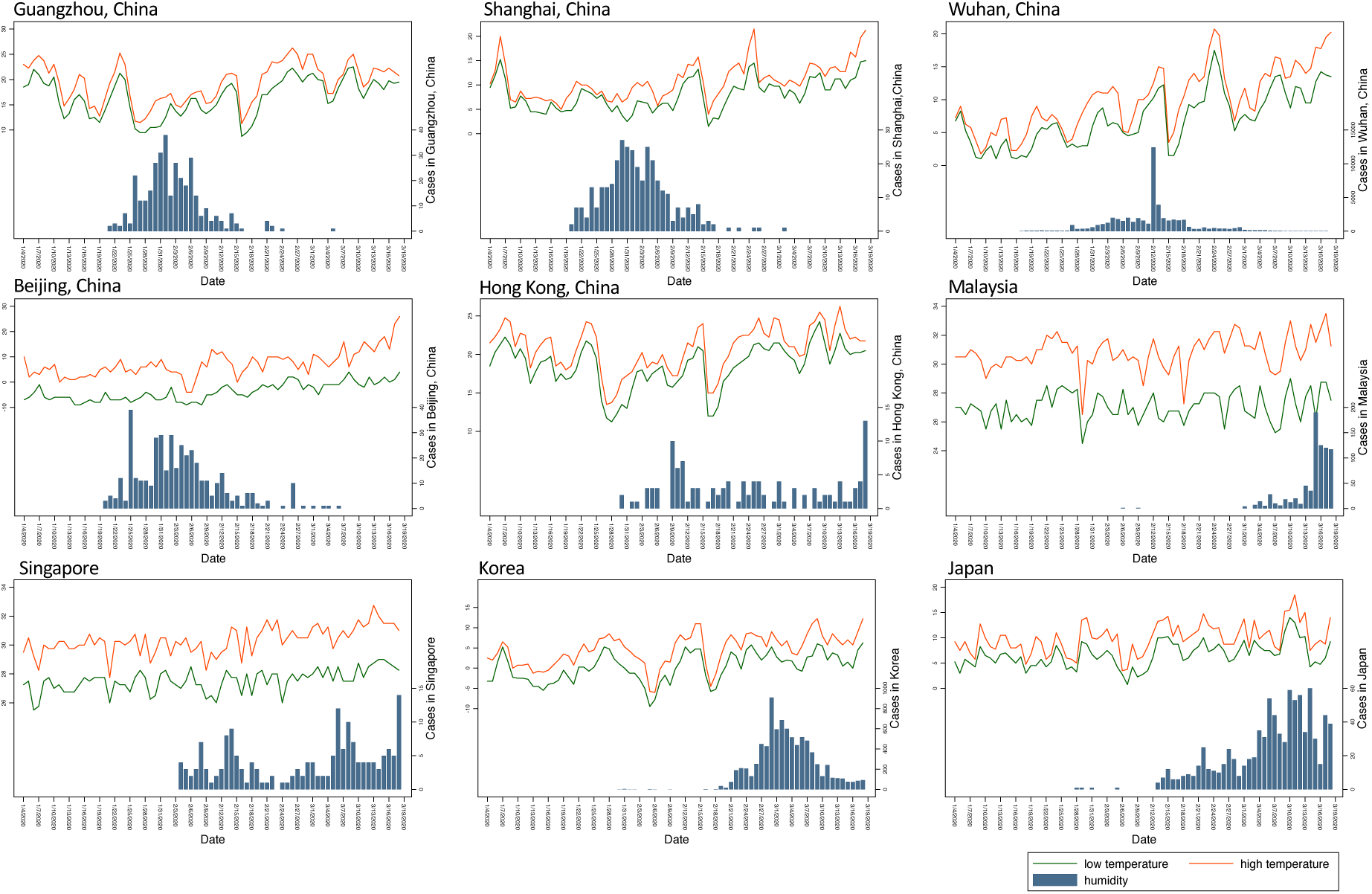
Daily temperature and distribution of COVID-19 daily new cases over time.

**Figure 2.**
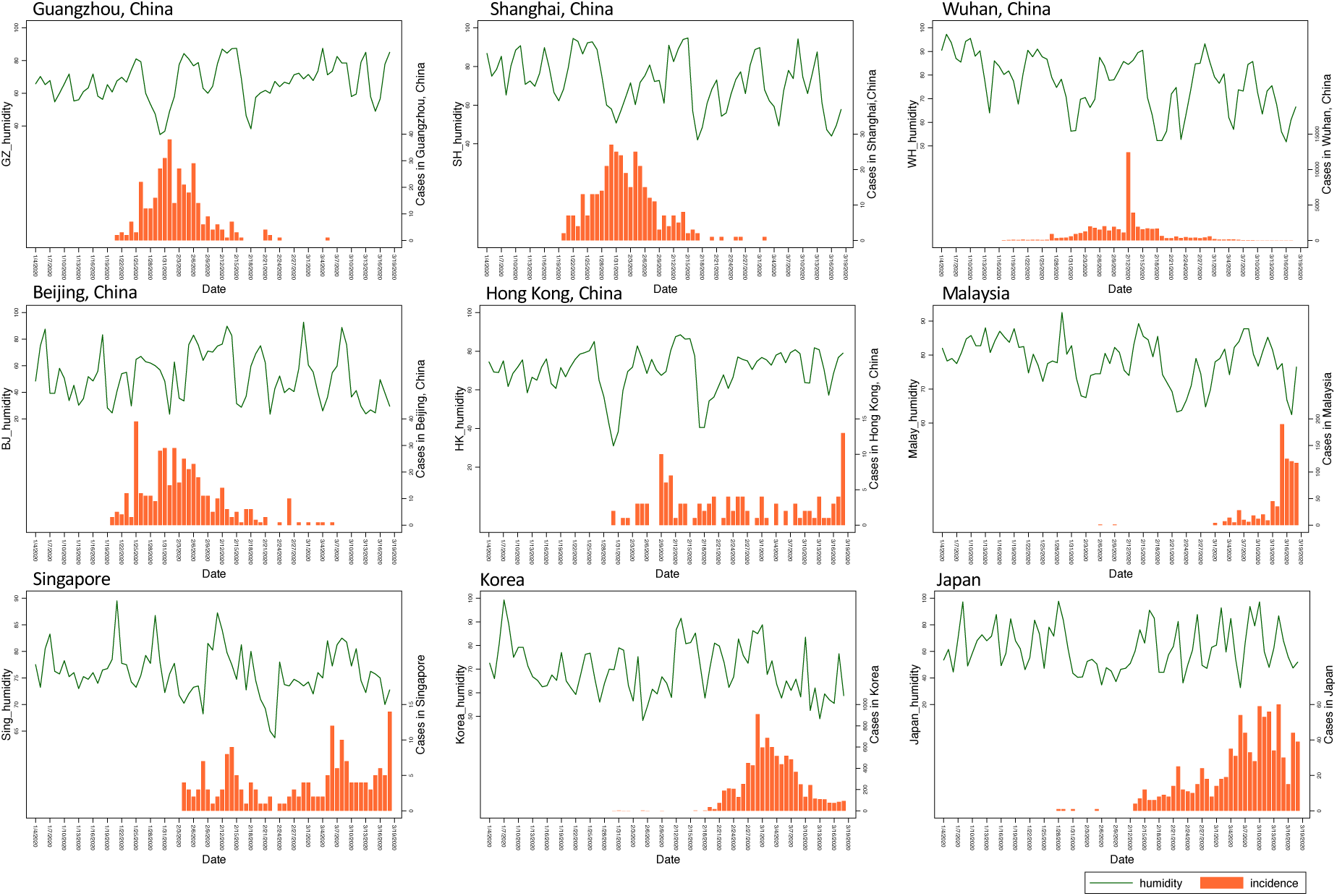
Daily relative humidity and distribution of COVID-19 daily new cases over time.

Pearson’s correlations between daily COVID-19 cases and meteorological factors are shown in Table 2. Daily confirmed new cases were negatively correlated with average temperature in Beijing (r = −0.565, P < 0.01), Shanghai (r = −0.471, P < 0.01), and Guangzhou (r = −0.530, P < 0.01), and, in contrast, positively correlated with that in Japan (r = 0.441, P < 0.01). The correlation between average temperature and relative humidity was found positive in Shanghai, Guangzhou, Hong Kong, Korea, and Japan, and negative in Beijing, Wuhan, Singapore, and Malaysia according to the pairwise Pearson correlation test (Table 2).

**Table 2.** Pearson correlation coefficient between daily COVID-19 new cases and meteorological factors.

| Variables | Daily new cases | P-value | Average temperature (°C) | P-value |
| --- | --- | --- | --- | --- |
| *Beijing, China* |  |  |  |  |
| (1) Daily new cases | 1.00 |  |  |  |
| (2) Average temperature (°C) | -0.57 | 0.00 | 1.00 |  |
| (3) Humidity (%) | 0.15 | 0.30 | -0.26 | 0.05 |
| *Shanghai, China* |  |  |  |  |
| (1) Daily new cases | 1.00 |  |  |  |
| (2) Average temperature (°C) | -0.47 | 0.00 | 1.00 |  |
| (3) Humidity (%) | -0.11 | 0.45 | 0.09 | 0.50 |
| *Wuhan, China* |  |  |  |  |
| (1) Daily new cases | 1.00 |  |  |  |
| (2) Average temperature (°C) | 0.04 | 0.81 | 1.00 |  |
| (3) Humidity (%) | 0.10 | 0.48 | -0.42 | 0.00 |
| *Guangzhou, China* |  |  |  |  |
| (1) Daily new cases | 1.00 |  |  |  |
| (2) Average temperature (°C) | -0.53 | 0.00 | 1.00 |  |
| (3) Humidity (%) | -0.29 | 0.05 | 0.34 | 0.01 |
| *Hong Kong, China* |  |  |  |  |
| (1) Daily new cases | 1.00 |  |  |  |
| (2) Average temperature (°C) | 0.08 | 0.56 | 1.00 |  |
| (3) Humidity (%) | 0.07 | 0.60 | 0.58 | 0.00 |
| *Singapore* |  |  |  |  |
| (1) Daily new cases | 1.00 |  |  |  |
| (2) Average temperature (°C) | 0.27 | 0.04 | 1.00 |  |
| (3) Humidity (%) | 0.04 | 0.74 | -0.58 | 0.00 |
| *Seoul, Korea* |  |  |  |  |
| (1) Daily new cases | 1.00 |  |  |  |
| (2) Average temperature (°C) | 0.29 | 0.02 | 1.00 |  |
| (3) Humidity (%) | 0.14 | 0.30 | 0.32 | 0.01 |
| *Tokyo, Japan* |  |  |  |  |
| (1) Daily new cases | 1.00 |  |  |  |
| (2) Average temperature (°C) | 0.42 | 0.00 | 1.00 |  |
| (3) Humidity (%) | 0.20 | 0.14 | 0.21 | 0.11 |
| *Kuala Lumpur, Malaysia* |  |  |  |  |
| (1) Daily new cases | 1.00 |  |  |  |
| (2) Average temperature (°C) | 0.21 | 0.11 | 1.00 |  |
| (3) Humidity (%) | -0.17 | 0.20 | -0.74 | 0.00 |

### GAM analysis of daily COVID-19 new cases with meteorological factors

The final GAM model of daily COVID-19 new cases incorporated date (time-series), average temperature, and mean relative humidity.

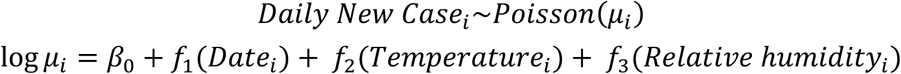

where the terms *f*_*1*_ to *f*_*2*_ are the smoothing functions, and β_0_ is the intercept. All estimates and significance levels were listed in Supplementary Table 1. The models with the best performance (lowest AIC) for each city is as follows: No-lag model for Shanghai and Singapore, Lag 1d model for Beijing and Wuhan, Lag 5d model for Guangzhou, Korea, and Kuala Lumpur, and Lag 14d model for Hong Kong and Japan (Table 3).

**Table 3.**
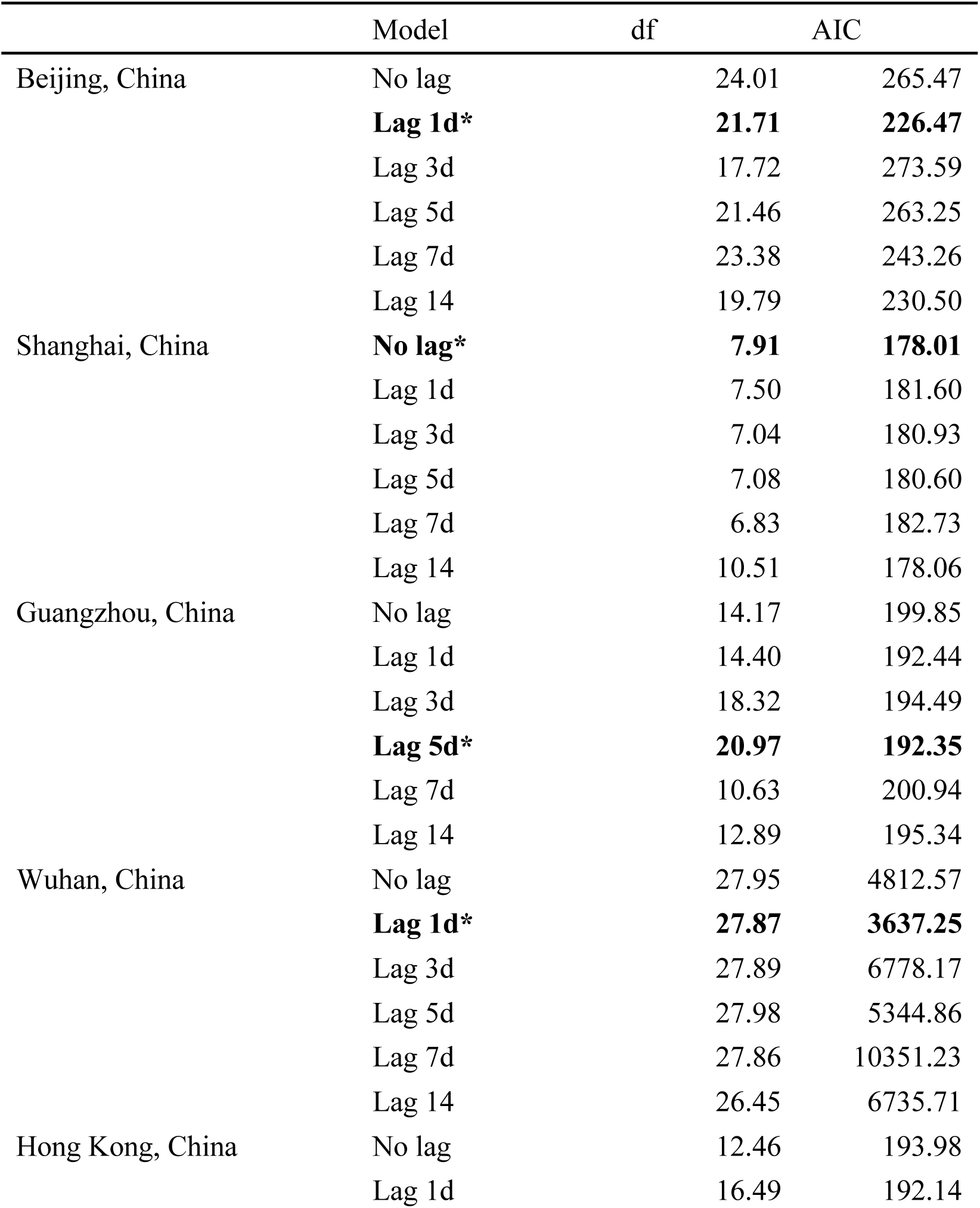
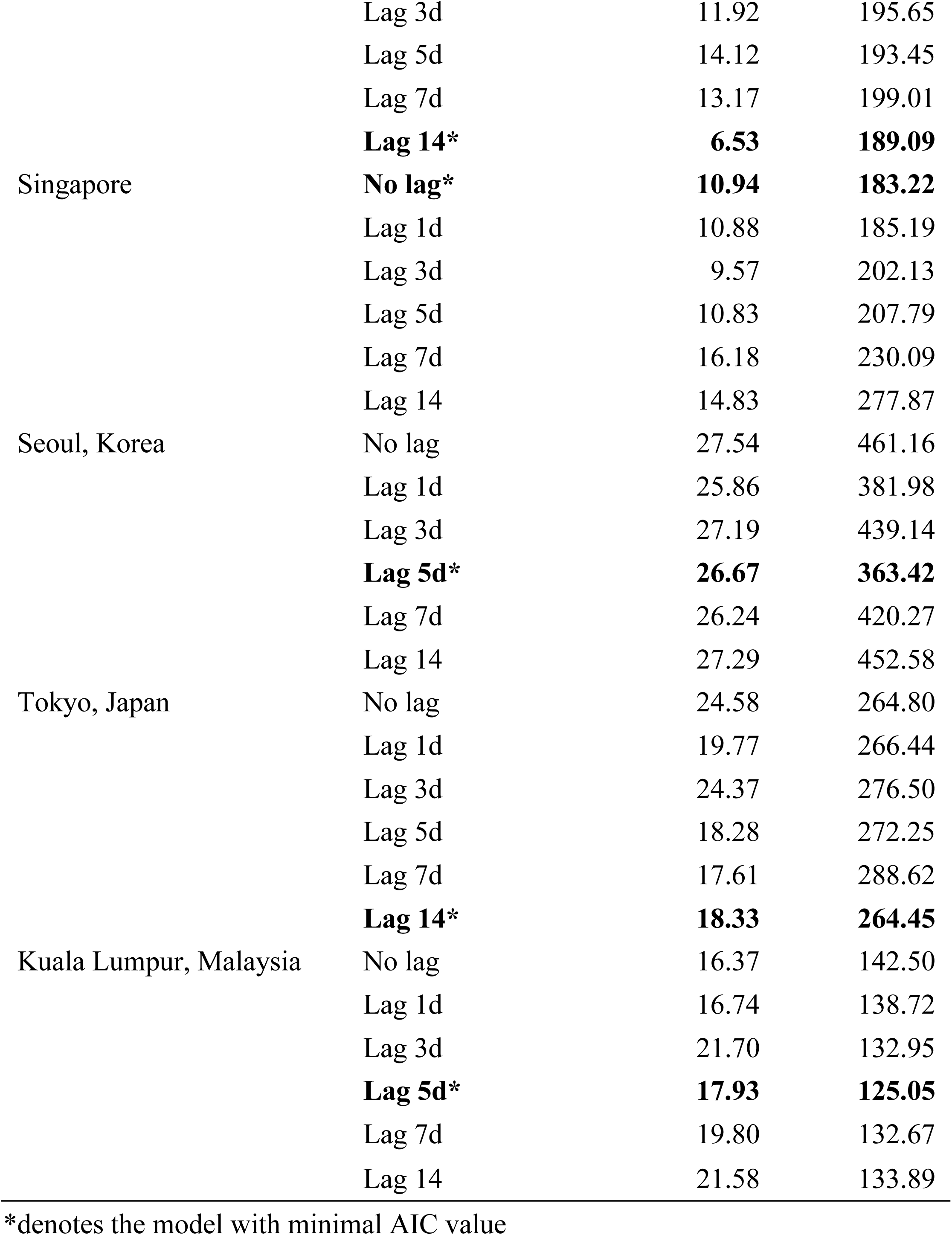
The selection of GAM model by Akaike’s An Information Criterion.

GAM results were reported using the smoothing components plot for temperature and relative humidity in Figures 3 and 4, respectively. The smoothing components plots demonstrated the estimated smoothing spline functions with the linear effect subtracted out, and each panel represented the weighted sum of basis functions for each time-varying covariate and is corresponding to the hypothesized model.

**Figure 3.**
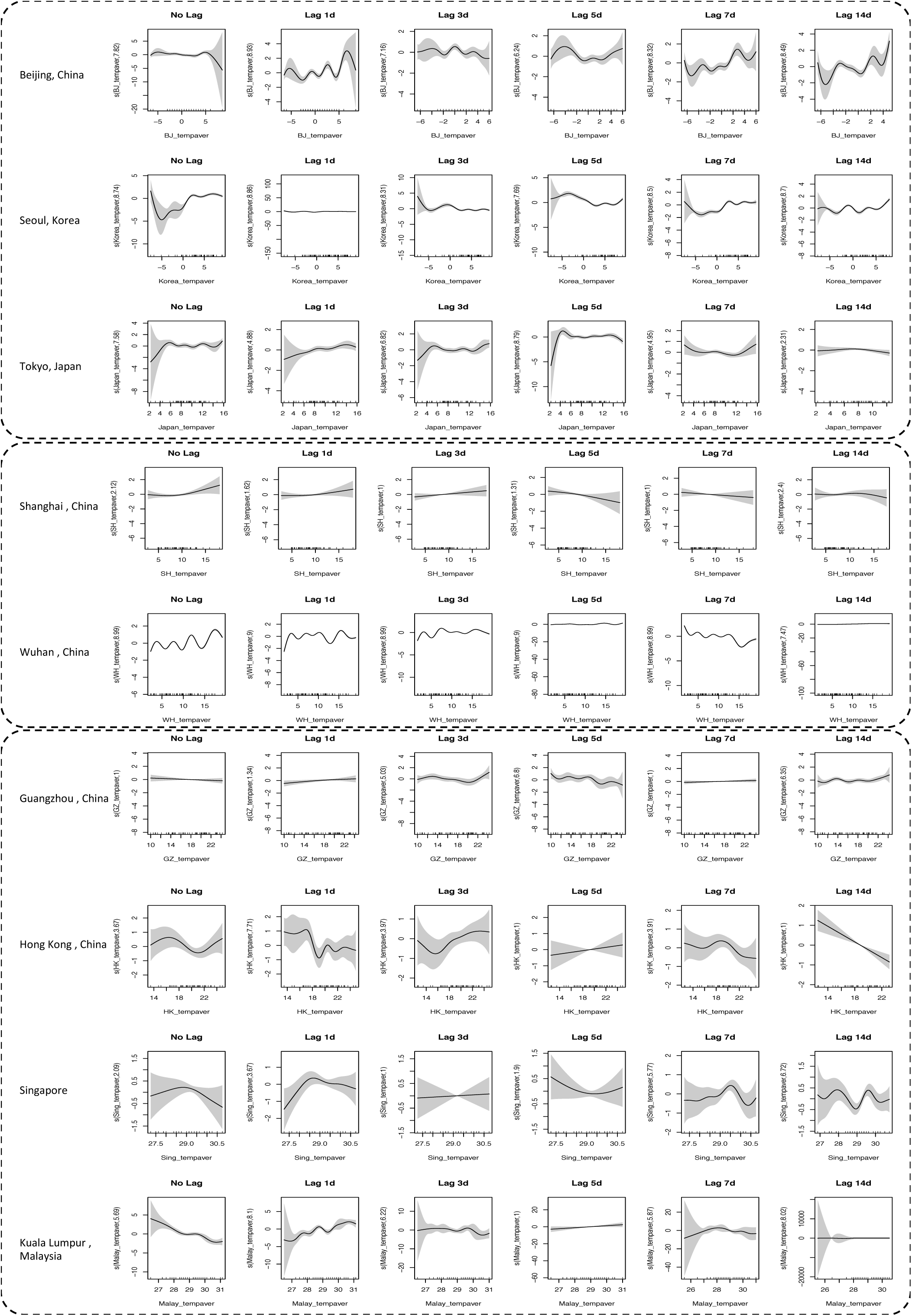
Smoothing component plots for daily COVID-19 new cases associated with average temperature.

**Figure 4.**
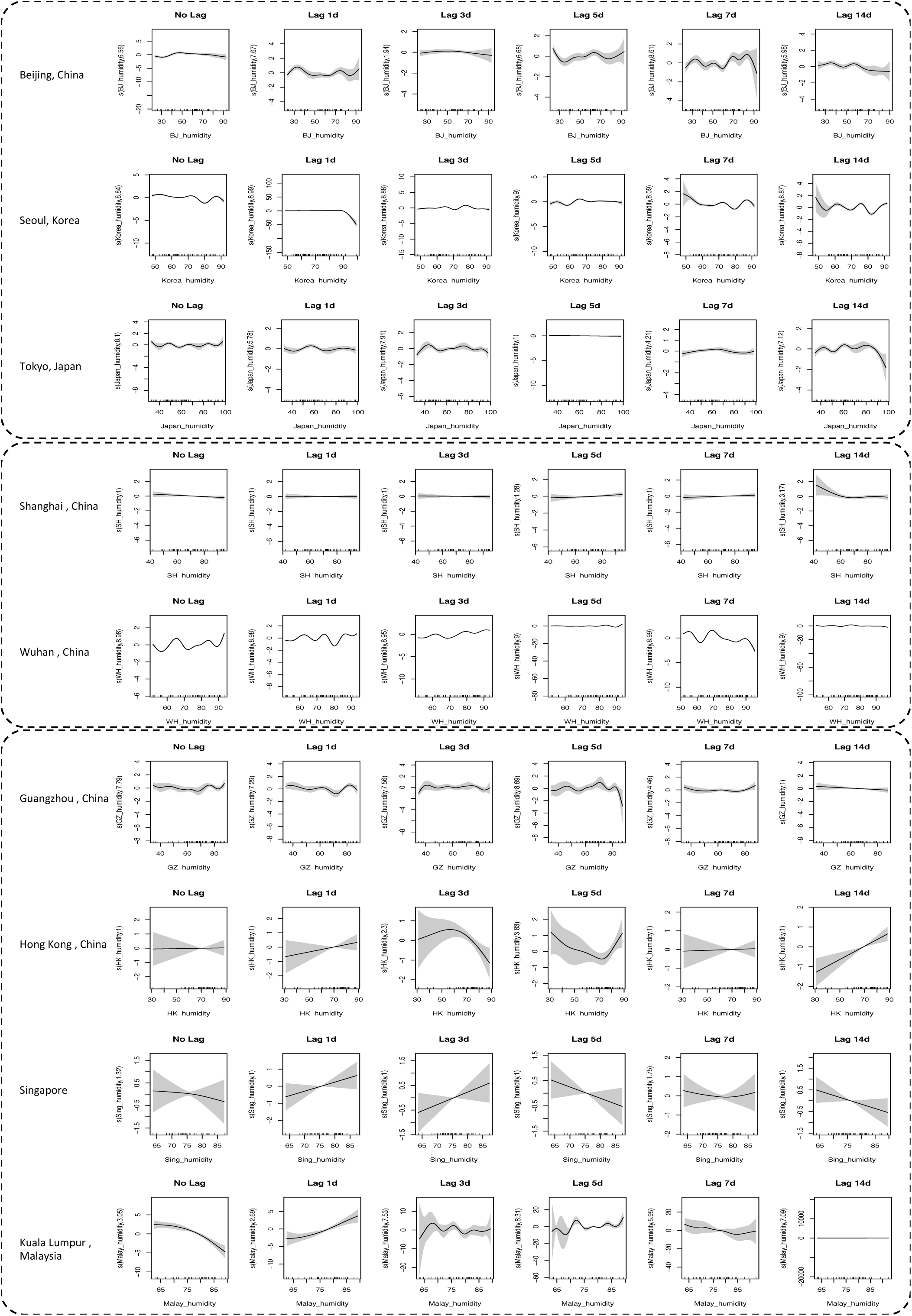
Smoothing component plots for daily COVID-19 new cases associated with average relative humidity.

The significant smoothers indicated that the correlations between new cases of COVID-19 and explanatory variables were non-linear. As shown in Figure 3, the no-lag model suggested that holding all linear and the other nonlinear terms fixed, daily new cases of COVID-19 tended to show no influence by the temperature, but the cases number decreased when temperature reached 5, 18, and 29, in Beijing, Hong Kong, and Singapore, respectively (P < 0.01 for all, Figure 3). While the magnitude of results might look small relative to the base rate of case accrual, these plots were on the log-case scale, so the effect on the case number is multiplicative. The distributions of COVID-19 cases displayed greater uncertainty at lower temperature in Beijing and Wuhan. Beijing and Wuhan has fluctuating patterns throughout the six models both regarding temperature and relative humidity except in lag 5d and lag 4d model for Wuhan, which might be due to the change of diagnostic method on February 12, when a total of 13,436 cases were added. Nevertheless, the relationship between relative humidity and new cases were less evident in the No-lag model, and the distributions of COVID-19 cases displayed greater uncertainty in Wuhan.

Nevertheless, in the Lag 1d model, distribution of COVID-19 in Beijing, Wuhan, Hong Kong and Malaysia revealed great uncertainty and high non-linearity. A hump of the increase was observed at around 28°C in Singapore. After the hump, the cases started to decrease in Singapore when temperature increases. A liner increase can be found in Guangzhou. Likewise, a significantly non-linear pattern was found in Wuhan between the relative humidity and COVID-19 cases. Figure 4 shows that during the study period, relative humidity has a positive effects for Singapore, Hong Kong as well as Malaysia constantly, with a linear pattern, yet a negative effects were discerned in Seoul when humidity greater than 90% in a non-linear way.

In Lag 3d model, the temperature has a positive influence on the daily new cases in Shanghai and Singapore in a linear pattern, and in Guangzhou when the temperature was greater than 20°C, in Hong Kong when temperature was greater than 15°C, and in Japan when temperature was between 2-4°C and 12-16°C in a non-linear pattern. Specifically, the distribution of the COVID-19 new cases in Beijing, Wuhan, and Kuala Lumpur was greatly fluctuating. Regarding the relative humidity, in Lag 3d model, Wuhan, Guangzhou, and Kuala Lumpur showed significant curvilinearity. A positive association between relative humidity and new cases can be found in Singapore, while a negative pattern was observed in Hong Kong when relative humidity greater than 65% (Figure 4).

In the Lag 5d model, COVID-19 new cases in Shanghai, Guangzhou, and Singapore when temperature less than 22°C were negatively correlated (Figure 3). A positive pattern can be discerned significantly in Tokyo when temperature was lower than 4°C, and in Hong Kong as well as in Kuala Lumpur. Regarding the relative humidity, except for Wuhan and Guangzhou that displayed highly fluctuating pattern, no evident effects could be discerned.

In the Lag 7d and Lag 14d models, Beijing, Wuhan, and Kuala Lumpur (only in Lag 14d model) showed great uncertainty of the relationship between temperature and daily new cases. Moreover, Shanghai and Singapore have different and inverse effects between Lag 7d and 14d models, where, in the Lag 7d model, Shanghai showed a negative pattern and Singapore showed a positive pattern while in the Lag 14d model, Shanghai showed a positive pattern and Singapore showed a negative pattern. Concerning the relative humidity, likewise, Beijing, Wuhan, and Guangzhou (only in Lag 14d model) displayed great uncertainty and curvilinearity (Figure 4). Hong Kong shows a positive pattern in both two model. Nevertheless, Shanghai and Singapore still showed inverse effects in the Lag 7d and 14d models, where in the Lag 7d model, both Shanghai and Singapore showed positive patterns while in the Lag 14d model, Shanghai and Singapore showed negative patterns.

## Discussion

In this study, we investigated the associations between meteorological factors and patterns of daily new cases of COVID-19 across nine Asian cities. Our analyses suggest a positive relationship between temperature and daily new cases of COVID-19 in Guangzhou, Singapore (except in the lagged 14d model), Hong Kong (except in the lagged 7d and 14d model) and Beijing (high curvilinearity). Relative humidity positively with the number of daily new cases in Singapore (except in the lagged 14d model), Hong Kong (except in the lagged 3d model). Negative association between relative humidity and the number of daily new cases was only observed in Shanghai consistently. Associations of temperature and relative humidity on the daily new cases were less evident in Tokyo, Seoul and Kuala Lumpur. The relationships in Wuhan presented a great fluctuating pattern. Therefore, our analysis suggest, unlike influenza, seasonality of COVID-19 may not be expected, and the epidemic is unlikely to fade out itself when summer comes.

Researchers have long been investigating how meteorological factors affect the viral infectivity, where the GAM has been frequently used, as it allows smooth components to be estimated for time, meteorological factors and other covariates, together with a non-smoothed period effect. Experiments in the last century reported that the influenza virus is more stable in cool and dry air (15, 16). And with increasing temperature, the viability of the influenza virus in aerosol or droplets(38), and the aerosol transmission diminishes (27). Lowen et al. reported that aerosol transmission of influenza between Guinea pigs was completely blocked at temperature higher than 30°C despite evidence of continuous viral shedding from infectious individuals; nevertheless, the direct contact transmission was not influenced, which was equally efficient at 30° C and 20° C (19). Chan et al. reported that the viability of SARS virus was rapidly lost (*>*3 log 10) at high temperatures (38^°^C) and high relative humidity (*>*95%)(31). The better stability of the SARS coronavirus at low temperatures and low humidity environment might facilitate its transmission in the community in subtropical areas (such as Hong Kong) during the spring and in air-conditioned environments. This might also explain why some Asian countries in tropical areas (such as Malaysia, Indonesia or Thailand) with high temperatures and high relative humidity environment did not have major community outbreaks of SARS. However, such an explanation is not currently convincing regarding the results in the present study, where the increase of the temperature increases the daily new cases of COVID-19 in some of the investigated communities. The high temperature and high relative humidity in the tropical Asian countries like Singapore, and Malaysia also seems to have little influence on the expanding daily new cases (Figure 3). However, this could have been confounded by multiple factors.

There are several reasons underlying the continuously expanding COVID-19 cases in Singapore and Malaysia, such as the high population density, massive gatherings, the use of air-conditioning and shortage of medical resources(31). The SARS-CoV-2 can persist at room temperature for up to 9 days, and its heat sensitivity rendered it susceptible to increased temperature, affecting its persistence in the outdoor environment. However, coronavirus was still found to be infective up to two weeks in the air-condition environment(31). As air-conditioners may increase the probability of spreading, it may be advisable to reduce the use of air-conditioners and keep ventilation. Moreover, during low temperature and high humidity, it is advisable to avoid massive gathering, as evidences are also supporting transmission by direct contact or close contact at the tropical area.

Humidity can influence aerosol transmission via altering the proportion of respiratory droplets undergoing aerosolization, and influencing the stability and viability of the virus within these aerosols. Respiratory droplets are generated in the high humidity of the respiratory tract. On entering an environment with low humidity, respiratory droplets reduce in size within seconds due to evaporation. At higher environmental humidity, respiratory droplets evaporate more slowly, and hence are larger and settle faster, and less aerosol nuclei are produced (39, 40). Previous studies have shown that influenza transmission between mice reduced as relative humidity increased from 47% to 70%(13, 41). Moreover, humidity can also influence indirect transmission by changing the mass of respiratory droplets accumulating on surfaces, and affecting the survival of the virus on surfaces. While increased humidity reduces the number of droplet nuclei formed, the same mechanisms (reduced droplet evaporation and faster droplet settling) result in a greater mass of respiratory droplets on surfaces (39, 40). Area with relatively low temperature and humidity has a higher infection rate comparing with tropical areas because the cold and dry weather is suitable for virus to survive and transmit (42). Viability of influenza virus appears greater at lower humidity, and progressively reduced influenza survival with increasing relative humidity over the range from 27% to 84%, with an increase in survival at 99% relative humidity, the mechanisms underlying which may be due to the low evaporation from droplets at high relative humidity maintain the solute concentration thus protecting the virus(27, 43). Alongside, 30-50^°^ N zone has become a zone for transmitting COVID-19 with similar average temperature of 5-11 °C and 47-79% humidity, which may be influenced by transoceanic migration of virus, but the underlying mechanism is still not understood (44).

Moreover, under laboratory conditions with constant humidity and temperature, circadian and circannual rhythms in variation of susceptibility of hosts (mice) have been observed(13, 45). Mice were substantially more susceptible to invasive pneumococcal disease in early morning hours than any other time of day(45), and more susceptible to influenza in winter than in summer (13) which was thought to be attributable to the daily and seasonal variation of melatonin (46). In addition, even in areas where many spend summers in air-conditioned spaces, marked annual variations in incidence constantly exist, and similar strains of virus appear almost simultaneously across vast stretches of oceans in areas of similar latitude around the globe(23, 24).

Therefore, although a higher temperature is associated with lower effectiveness of virus transmission, yet it does not necessarily suggest a lower chance for virus to survive. The natural fading out of the virus in summer is unlikely given the widespread use of air conditioners in developed areas and the dense population in the city area. Until an effective vaccine becomes widely available for the establishment of herd immunity and the discovery of efficient pharmaceutical therapies, strict public-health measures should be implemented, including social distancing, quarantine, tracing, facemask wearing, and promotion of hand washing.

## Strength and limitations

This is the first study to statistically analyze the relationship between meteorological factors and the daily new cases of COVID-19. Because of the non-linear nature of the data, we also performed GAMs to quantitate and visualize the relationship using splines functions. However, there are several limitations. Firstly, numbers of cases in Malaysia, Korea, and Japan were obtained from the epidemiologic reports released by the Department of Health in the corresponding countries, instead of a daily update of case numbers, which is not available for these countries. Hence, some cases might be missing. Secondly, the duration of the study period was short and the number of cases in Malaysia, Singapore and Japan were small. Thirdly, we only considered two meteorological factors (temperature and humidity) in this study. Other covariates such as wind speed, pollutant concentration, population density, air conditioning use, rainfall, which could also influence the spread of COVID-19, were not included. Moreover, while we collected the meteorological data from the capital cities of Malaysia, Japan, and Korea, the number of domestic cases at the national level in these three countries was used for analysis due to lack of available data at their city level. Most importantly, we did not incorporate the public health measures into the modelling, which may greatly confound the results, but we chose the date when accumulated confirmed cases exceeding 30, based on the postulation that a certain level of public health measures had been carried out at that time, thus the confounding effects can be mitigated at some point.

## Conclusion

In this study, we found inconsistent associations between temperature and relative humidity and the number of COVID-19 daily cases across cities and time. Therefore, unlike influenza, seasonality of COVID-19 may not be expected, and the epidemic is unlikely to fade out when summer arrives. Strict public health measures such as social distancing, quarantine, tracing, facemask wearing, and promotion of hand washing are needed until a vaccine becomes widely available and induce the herd immunity among the community.

## Data Availability

Open-source data were used.

## Supplementary materials

**Supplementary Table 1.**
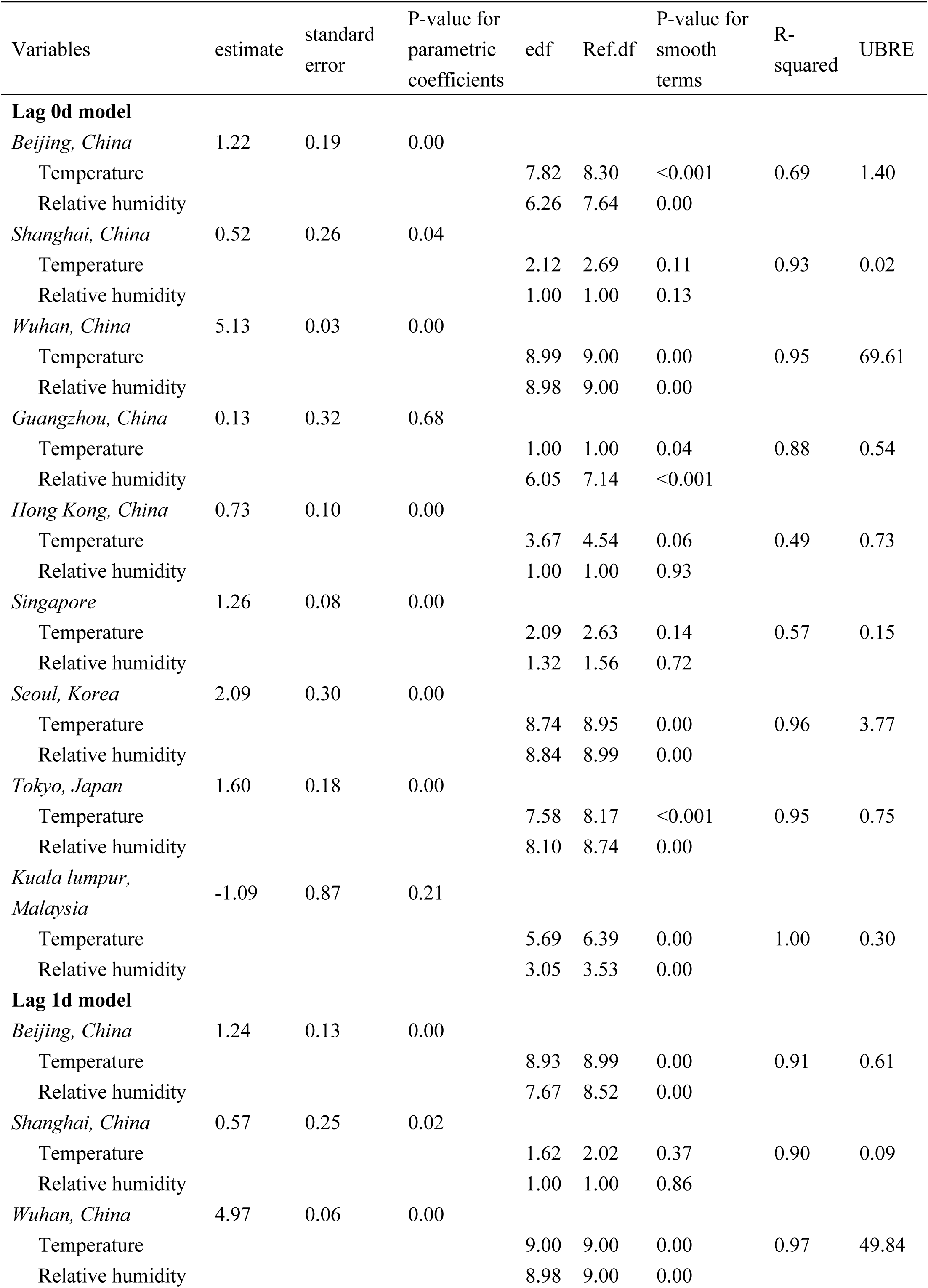
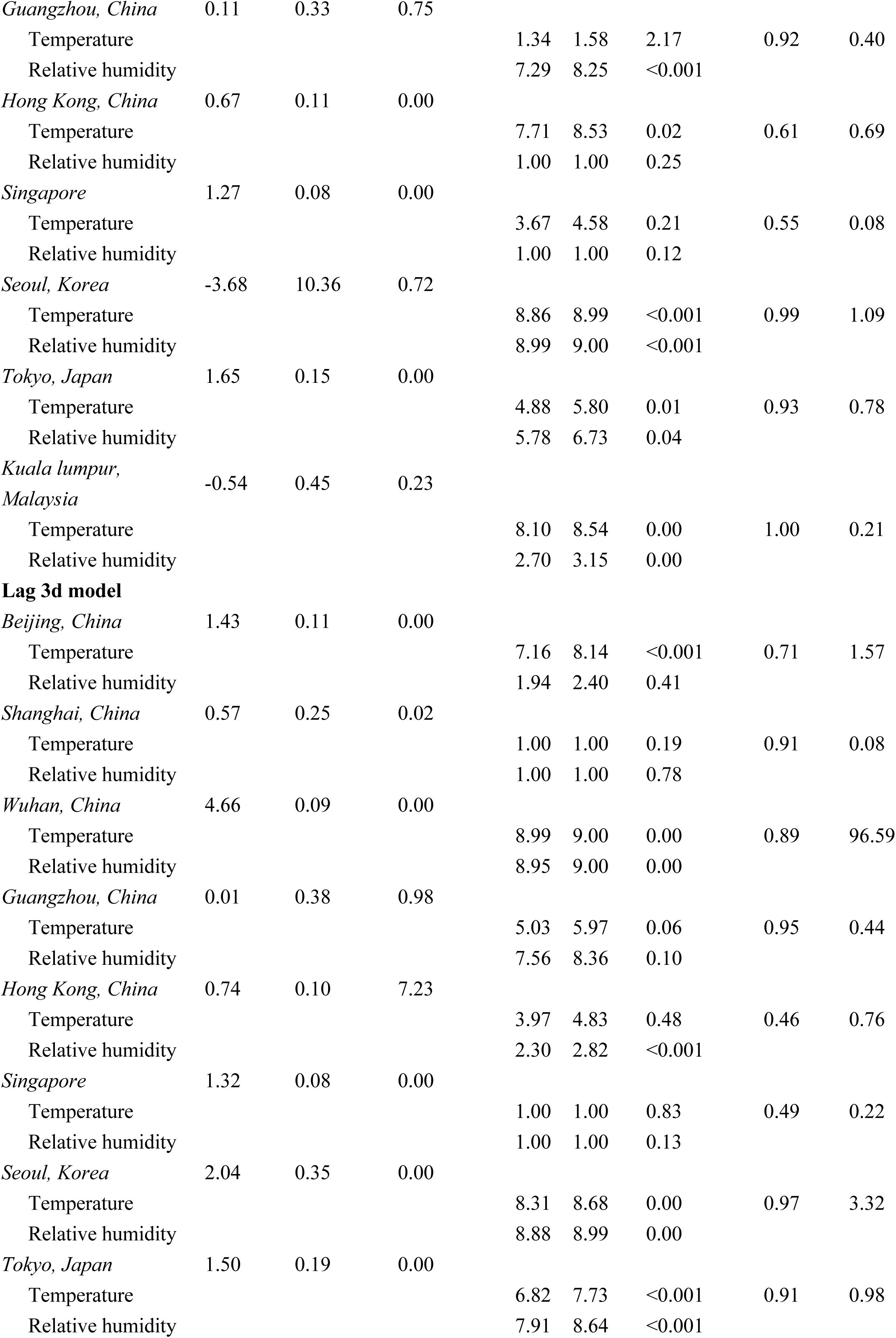
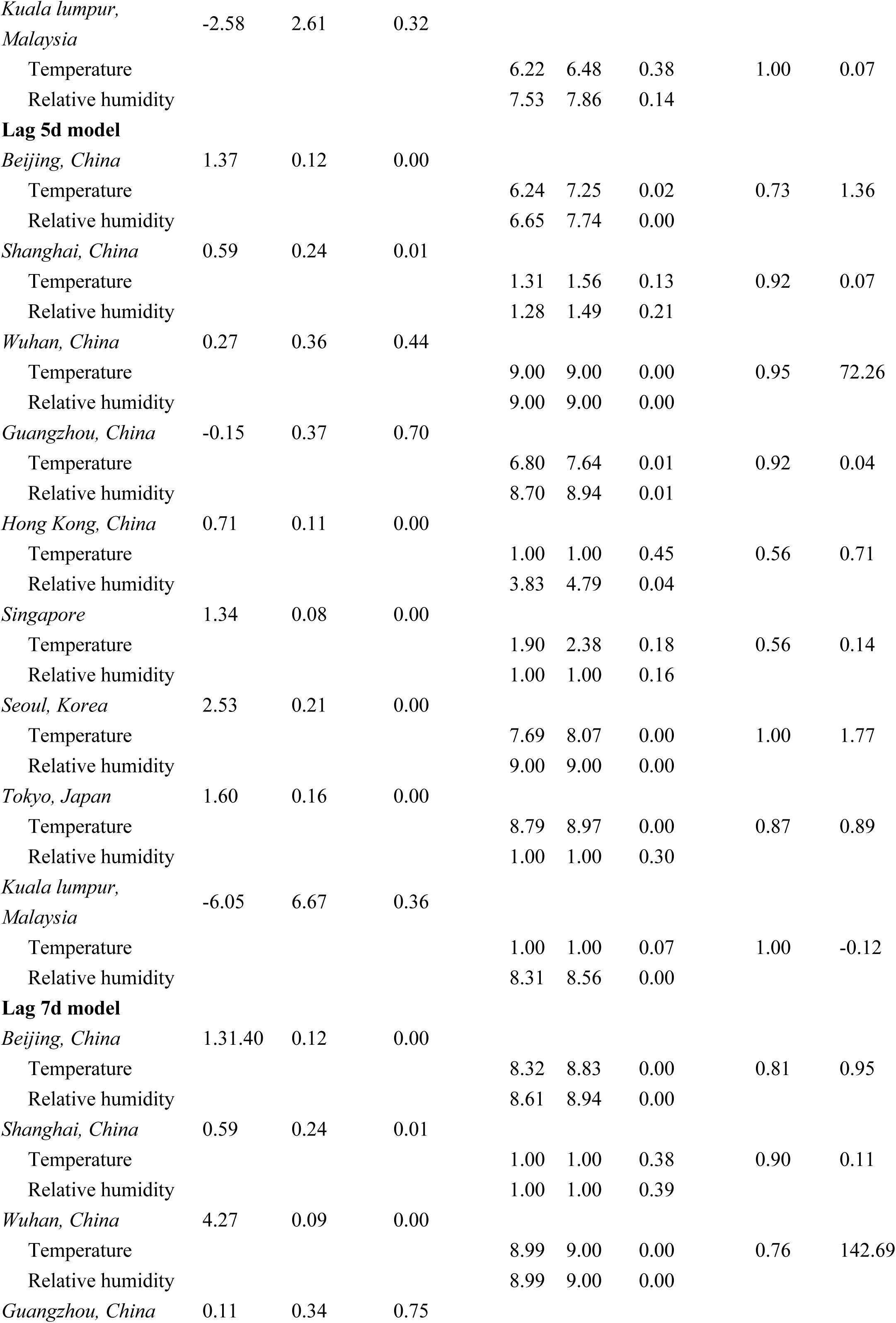
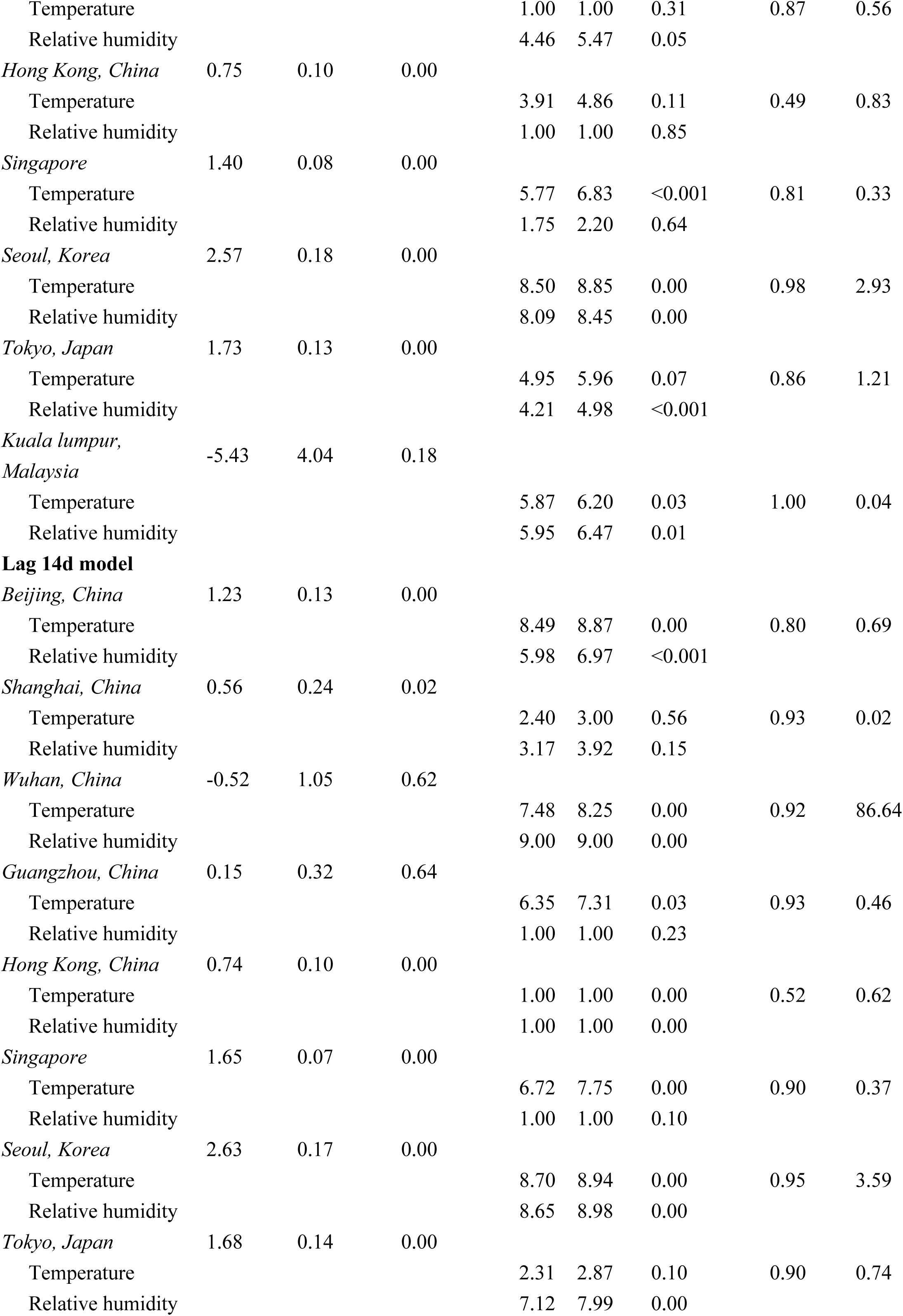
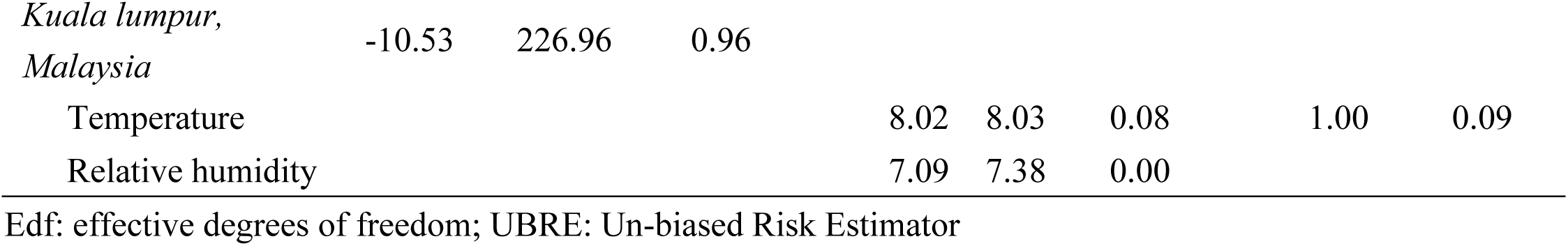
The effects of temperature and relative humidity on daily new cases of COVID-19.

## Reference

1. Chen N, Zhou M, Dong X, Qu J, Gong F, Han Y, et al. Epidemiological and clinical characteristics of 99 cases of 2019 novel coronavirus pneumonia in Wuhan, China: a descriptive study. 2020.

2. The World Health Organization. Statement on the second meeting of the International Health Regulations (2005) Emergency Committee regarding the outbreak of novel coronavirus (2019-nCoV). [Available 30 Jan 2020, cited 24 Feb 2020], Available from: https://www.who.int/news-room/detail/30-01-2020-statement-on-the-second-meeting-of-the-international-health-regulations-(2005)-emergency-committee-regarding-the-outbreak-of-novel-coronavirus-(2019-ncov).

3. Xu X, Chen P, Wang J, Feng J, Zhou H, Li X, et al. Evolution of the novel coronavirus from the ongoing Wuhan outbreak and modeling of its spike protein for risk of human transmission. 2020:1–4.

4. Gralinski LE, Menachery VDJV. Return of the Coronavirus: 2019-nCoV. 2020;12(2):135.

5. Cowling BJ, Leung GM. Epidemiological research priorities for public health control of the ongoing global novel coronavirus (2019-nCoV) outbreak. 2020.

6. Tang J. The emergence and spread of the 2019 novel coronavirus (2019-nCoV).

7. Riou J, Althaus CLJE. Pattern of early human-to-human transmission of Wuhan 2019 novel coronavirus (2019-nCoV), December 2019 to January 2020. 2020;25(4).

8. Lauer SA, Grantz KH, Bi Q, Jones FK, Zheng Q, Meredith HR, et al. The incubation period of coronavirus disease 2019 (COVID-19) from publicly reported confirmed cases: Estimation and application. 2020.

9. Lai C-C, Shih T-P, Ko W-C, Tang H-J, Hsueh P-RJIjoaa. Severe acute respiratory syndrome coronavirus (SARS-CoV-2) and corona virus disease-2019 (COVID-19): the epidemic and the challenges. 2020:105924.

10. WHO. Coronavirus disease 2019 (COVID-19) Situation Report – 68. [published 28 March 2020, cited 29 March 2020], Available from: https://www.who.int/docs/default-source/coronaviruse/situation-reports/20200328-sitrep-68-covid-19.pdf?sfvrsn=384bc74c_2

11. de Wit E, van Doremalen N, Falzarano D, Munster VJJNRM. SARS and MERS: recent insights into emerging coronaviruses. 2016;14(8):523.

12. Cai Q-C, Lu J, Xu Q-F, Guo Q, Xu D-Z, Sun Q-W, et al. Influence of meteorological factors and air pollution on the outbreak of severe acute respiratory syndrome. 2007;121(4):258–65.

13. Schulman JL, Kilbourne EDJJoEM. Experimental transmission of influenza virus infection in mice: II. Some factors affecting the incidence of transmitted infection. 1963;118(2):267–75.

14. Horby P, Nguyen NY, Dunstan SJ, Baillie JKJPo. The role of host genetics in susceptibility to influenza: a systematic review. 2012;7(3).

15. Hemmes J, Winkler K, Kool SJN. Virus survival as a seasonal factor in influenza and poliomyelitis. 1960;188(4748):430–1.

16. Loosli C, Lemon H, Robertson O, Appel EJPotSfEB, Medicine. Experimental Air-Borne Influenza Infection. I. Influence of Humidity on Survival of Virus in Air. 1943;53(2):205–6.

17. Paynter S. Humidity and respiratory virus transmission in tropical and temperate settings. Epidemiology and infection. 2015;143(6):1110–8.

18. Finkelman BS, Viboud C, Koelle K, Ferrari MJ, Bharti N, Grenfell BTJPo. Global patterns in seasonal activity of influenza A/H3N2, A/H1N1, and B from 1997 to 2005: viral coexistence and latitudinal gradients. 2007;2(12).

19. Lowen AC, Steel J, Mubareka S, Palese PJJov. High temperature (30 C) blocks aerosol but not contact transmission of influenza virus. 2008;82(11):5650–2.

20. Organization WH. Coronavirus disease 2019 (COVID-19): situation report, 28. 2020.

21. Addie D, Schaap I, Nicolson L, Jarrett OJJogv. Persistence and transmission of natural type I feline coronavirus infection. 2003;84(10):2735–44.

22. Arbour N, Côté G, Lachance C, Tardieu M, Cashman NR, Talbot PJJJov. Acute and persistent infection of human neural cell lines by human coronavirus OC43. 1999;73(4):3338–50.

23. Langmuir AD, Schoenbaum SCJHp. The epidemiology of influenza. 1976;11(10):49–56.

24. THACKER Sbjer. The persistence of influenza A in human populations. 1986;8(1):129–42.

25. Hope-Simpson R, Golubev DJE, Infection. A new concept of the epidemic process of influenza A virus. 1987;99(1):5–54.

26. Hammond G, Raddatz R, Gelskey DJRoid. Impact of atmospheric dispersion and transport of viral aerosols on the epidemiology of influenza. 1989;11(3):494–7.

27. Paynter SJE, Infection. Humidity and respiratory virus transmission in tropical and temperate settings. 2015;143(6):1110–8.

28. Tamerius J, Nelson MI, Zhou SZ, Viboud C, Miller MA, Alonso WJJEhp. Global influenza seasonality: reconciling patterns across temperate and tropical regions. 2011;119(4):439–45.

29. Lofgren E, Fefferman NH, Naumov YN, Gorski J, Naumova ENJJov. Influenza seasonality: underlying causes and modeling theories. 2007;81(11):5429–36.

30. Tamerius JD, Shaman J, Alonso WJ, Bloom-Feshbach K, Uejio CK, Comrie A, et al. Environmental predictors of seasonal influenza epidemics across temperate and tropical climates. 2013;9(3).

31. Chan K, Peiris J, Lam S, Poon L, Yuen K, Seto WJAiv. The effects of temperature and relative humidity on the viability of the SARS coronavirus. 2011;2011.

32. Hastie T, Tibshirani RJSmimr. Generalized additive models for medical research. 1995;4(3):187–96.

33. Katsouyanni K, Touloumi G, Samoli E, Gryparis A, Le Tertre A, Monopolis Y, et al. Confounding and effect modification in the short-term effects of ambient particles on total mortality: results from 29 European. cities within the APHEA2 project. 2001:521–31.

34. Peng RD, Dominici F, Louis TAJJotRSSSA. Model choice in time series studies of air pollution and mortality. 2006;169(2):179–203.

35. Kan H, London SJ, Chen G, Zhang Y, Song G, Zhao N, et al. Differentiating the effects of fine and coarse particles on daily mortality in Shanghai, China. 2007;33(3):376–84.

36. Huang Y, Deng T, Yu S, Gu J, Huang C, Xiao G, et al. Effect of meteorological variables on the incidence of hand, foot, and mouth disease in children: a time-series analysis in Guangzhou, China. 2013;13(1):134.

37. Huang C, Wang Y, Li X, Ren L, Zhao J, Hu Y, et al. Clinical features of patients infected with 2019 novel coronavirus in Wuhan, China. 2020.

38. Dublineau A, Batejat C, Pinon A, Burguiere AM, Manuguerra J-CJPO. Persistence of the 2009 pandemic influenza A (H1N1) virus in water and on non-porous surface. 2011;6(11).

39. Xie X, Li Y, Chwang A, Ho P, Seto WJIa. How far droplets can move in indoor environments--revisiting the Wells evaporation-falling curve. 2007;17(3):211–25.

40. Yang W, Marr LCJPo. Dynamics of airborne influenza A viruses indoors and dependence on humidity. 2011;6(6).

41. Schulman JL, Kilbourne EDJN. Airborne transmission of influenza virus infection in mice. 1962;195(4846):1129–30.

42. To K, Lo AWJTJoPAJotPSoGB, Ireland. Exploring the pathogenesis of severe acute respiratory syndrome (SARS): the tissue distribution of the coronavirus (SARS-CoV) and its putative receptor, angiotensin-converting enzyme 2 (ACE2). 2004;203(3):740–3.

43. Yang W, Elankumaran S, Marr LCJPo. Relationship between humidity and influenza A viability in droplets and implications for influenza’s seasonality. 2012;7(10).

44. Sajadi MM, Habibzadeh P, Vintzileos A, Shokouhi S, Miralles-Wilhelm F, Amoroso AJAaS. Temperature and Latitude Analysis to Predict Potential Spread and Seasonality for COVID-19. 2020.

45. Feigin RD, San Joaquin VH, Haymond MW, Wyatt Rgjn. Daily periodicity of susceptibility of mice to pneumococcal infection. 1969;224(5217):379–80.

46. Nelson RJ, Drazen DLJRND. Melatonin mediates seasonal adjustments in immune function. 1999;39(3):383–98.

